## Supplementary files for "Serology assays used in SARS-CoV-2 seroprevalence surveys worldwide: a systematic review and meta-analysis of assay features, testing algorithms, and performance"

**Supplementary file 1**

### **Figure S1. PRISMA diagram**

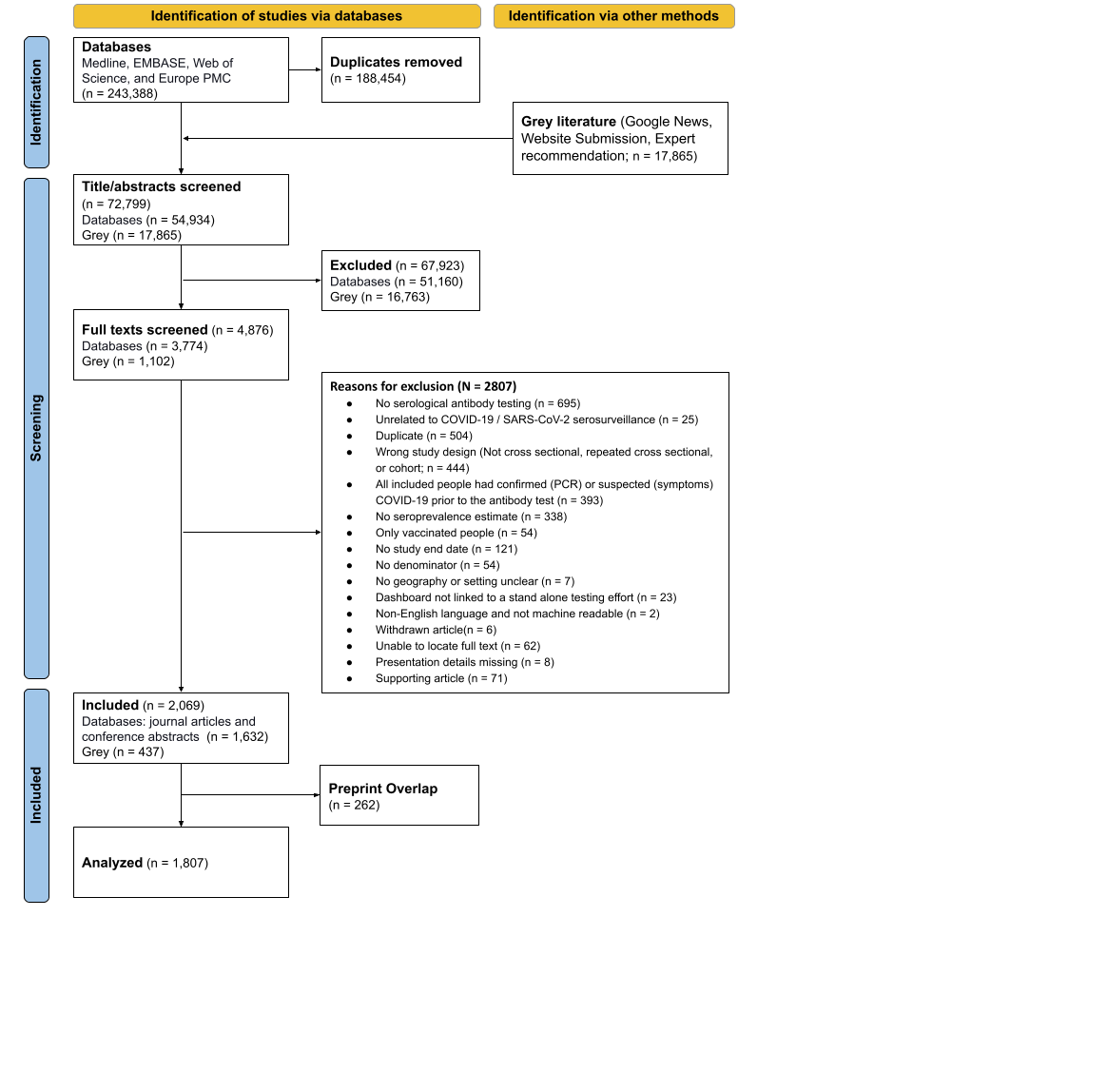

### **Table S1. The major categories of assay**

| Categories of assay | Description |
| --- | --- |
| Neutralization assays | Neutralization assays are lab-based tests that use live or pseudotyped viruses to determine if patient antibodies can prevent viral infection in vitro. |
| Lateral flow immunoassays (LFIAs) | LFIA technology is a type of rapid test, which is easily administered at the point of care by minimally trained personnel and at low cost, administered self with a rapid time-to-result turnaround. |
| Immunofluorescence assays (IFAs) | In IFAs, antibodies bind to the SARS-CoV-2 protein of interest. Fluorescent dyes are coupled to these immune complexes in order to visualize the protein of interest using microscopy or flow cytometry. |
| Enzyme-linked immunosorbent assays (ELISAs) | ELISA assay, also a lab-based test, is the most commonly used format of the serological test. Wells are coated with the viral antigen of interest. After the formation of immune complexes, an enzyme-linked secondary antibody followed by the enzyme’s substrate is added, which results in chemical modification of the substrate in the presence of immune complexes, which is measured colorimetrically through spectroscopy. The relative light absorption at a particular wavelength is directly proportional to the number of labeled complexes present. |
| Chemiluminescence serology assays (CLIAs) | CLIA technology follows a similar concept to ELISA by taking advantage of high binding affinity between the viral antigen(s) and host antibodies but uses chemical probes that yield light responses that are directly proportional to the amount of labeled immune complexes present. |
| Other | Assays do not fall into the above five categories. |

### **Table S2. Reference panels of five third-party lab evaluations**

| **Third Party** | **Country** | **Time of Evaluation** | **Type of Assay** | **Isotype Detected** | **Overall Sample Size** | **Positive Sample** | **Negative Sample** | **Analysis** |
| --- | --- | --- | --- | --- | --- | --- | --- | --- |
| **FDA [1]** | USA | Since 2020 (Ongoing) | All types (including LFIAs, IFAs, ELISAs, CLIAs, and others, n = NA) | IgM, IgG, IgA, and total antibody | n >= 120 | Confirmed with NAAT; disease severity unknown; time PSO unknown (*n* = 30) | Pre-pandemic negative serum and plasma (prior to 2020) without regard for clinical status (*n*=70); HIV+ (*n*=10) | Sensitivity calculated as the fraction of positive results among true positives, and specificity as the fraction of negative results among true negatives. Estimates were stratified by isotype (e.g., IgM, IgG, IgA, and total antibody) separately and combined where available. |
| **FIND Diagnostics [2,3]** | France, USA, Italy, Brazil, Spain, UK, Switzerland | March 2020 - March 2021 | LFIAs (n=35) and ELISAs (n=16) | IgM, IgG, IgA, and total antibody | 199 - 461 | Confirmed with RT-PCR (*n* > 100); disease severity varies; time PSO (*n* = 10 for day 0-3, *n* = 20 for day 4-7, *n* = 30 for day 8-14, *n* = 20 for day 15-28, *n* = 20 for day 29+ by design but varied between site) | Pre-pandemic samples (prior to 2018; target *n* > 300) | Sensitivity calculated as the fraction of positive results among true positives, and specificity as the fraction of negative results among true negatives along with GLM fit with site and time PSO included as random effects. Estimates were stratified by time PSO, isotype (e.g., IgM, IgG, IgA, and total antibody) separately and combined where available. |
| **Netherland CIDC [4]** | Netherland | July 2020 | LFIAs, IFAs, ELISAs, and CLIAs (n=47) | IgM, IgG, and total antibody | 9 - 489 | Confirmed with RT-PCR; disease severity varies; time PSO stratified by before or after 14 days (*n* varies between test and site) | Pre-pandemic samples (prior to Dec 2019); syndromic patients with potentially cross-reactive microorganisms during pandemic (*n* varies between test and site) | Sensitivity calculated as the fraction of positive results among true positives, and specificity as the fraction of negative results among true negatives (95% CI based on Wilson score). Sensitivity and specificity were stratified by disease severity, time PSO, isotype (e.g., IgM, IgG, IgA, and total antibody) separately and combined where available. |
| **Doherty Institute [5,6]** | Australia | April 2020 - April 2021 | LFIA (n=27) | IgM, IgG, and total antibody | Varied in each phase of evaluation | Confirmed with RT-PCR; disease severity unknown; time PSO varies (*n =* 50-100, varies between the phase of evaluation) | Pre-pandemic samples (2018 and 2019) and non-COVID 19 infections (*n* = 92-100); cross-reactivity panel (*n* = 25; malaria, Influenza A, Influenza B, CMV IgM positive, acute parvovirus B19, EBV, mycoplasma, parainfluenza, C. psittaci, toxoplasma, rubella, and Hepatitis A and B) | Sensitivity calculated as the fraction of positive results among true positives, and specificity as the fraction of negative results among true negatives. Estimates were stratified by time PSO, isotype (e.g., IgM, IgG, and total antibody) separately and combined where available. |
| **NRL-WHO [7]** | Australia | Since December 2020 (Ongoing) | LFIAs, ELISAs, and CLIAs (n = 40) | IgG, IgM | NA | Confirmed with NAAT or clinical diagnosis; disease severity unknown; time PSO >14 days or >14 days post-NAAT (*n* unknown) | Pre-pandemic samples (prior to Nov 2019) (*n* unknown); cross-reactivity panel (*n* = 55; CMB IgM, EBV VCA IgM, Influenza A, Influenza B, Hepatitis A, Hepatitis B, Hepatitis C, HIV, malaria, mycoplasma, parainfluenza, parvovirus, C. psittaci, rubella, syphilis, toxoplasma) | Sensitivity calculated as the fraction of positive results among true positives, and specificity as the fraction of negative results among true negatives. Estimates were stratified by time PSO, isotype (e.g., IgM, IgG, and total antibody) separately and combined where available. |

### **Table S3. Testing characteristics of included seroprevalence studies**

| **Study Characteristics** | **Total** | | **Africa** | | **Americas** | | **Eastern Mediterranean** | | **Europe** | | **South-East Asia** | | **Western Pacific** | |
| --- | --- | --- | --- | --- | --- | --- | --- | --- | --- | --- | --- | --- | --- | --- |
|  | **(N = 1807)** | | **(N = 89)** | | **(N = 522)** | | **(N = 98)** | | **(N = 846)** | | **(N = 129)** | | **(N = 123)** | |
|  | **n** | **%** | **n** | **%** | **n** | **%** | **n** | **%** | **n** | **%** | **n** | **%** | **n** | **%** |
| Sample Size (Median [IQR]) | 1240  (416-4558) | | 1769  (660-4258) | | 1096  (388-4445) | | 842  (388-2244) | | 1214  (386-4549) | | 1880  (728-10310) | | 1228  (512-6929) | |
| Sampling End Date^a^ |  |  |  |  |  |  |  |  |  |  |  |  |  |  |
| *2020 Q1-Q2* | 668 | 37.0 | 12 | 13.5 | 187 | 35.8 | 32 | 32.7 | 361 | 42.7 | 11 | 8.5 | 65 | 52.8 |
| *2020 Q3-Q4* | 862 | 47.7 | 61 | 68.5 | 275 | 52.7 | 53 | 54.1 | 344 | 40.7 | 82 | 63.6 | 47 | 38.2 |
| *2021 Q1-Q2* | 248 | 13.7 | 16 | 18.0 | 55 | 10.5 | 12 | 12.2 | 128 | 15.1 | 27 | 20.9 | 10 | 8.1 |
| *2021 Q3-Q4* | 29 | 1.6 | 0 | 0.0 | 5 | 1.0 | 1 | 1.0 | 13 | 1.5 | 9 | 7.0 | 1 | 0.8 |
| Specimen Type |  |  |  |  |  |  |  |  |  |  |  |  |  |  |
| *Serum* | 1016 | 56.2 | 34 | 38.2 | 257 | 49.2 | 66 | 67.3 | 489 | 57.8 | 82 | 63.6 | 88 | 71.5 |
| *Plasma* | 112 | 6.2 | 10 | 11.2 | 38 | 7.3 | 7 | 7.1 | 47 | 5.6 | 7 | 5.4 | 3 | 2.4 |
| *Whole blood* | 255 | 14.1 | 39 | 43.8 | 72 | 13.8 | 13 | 13.3 | 108 | 12.8 | 10 | 7.8 | 13 | 10.6 |
| *Dried blood* | 43 | 2.4 | 3 | 3.4 | 24 | 4.6 | 0 | 0.0 | 16 | 1.9 | 0 | 0.0 | 0 | 0.0 |
| *Multiple types* | 93 | 5.1 | 1 | 1.1 | 29 | 5.6 | 4 | 4.1 | 51 | 6.0 | 6 | 4.7 | 2 | 1.6 |
| *Not reported* | 288 | 15.9 | 2 | 2.2 | 102 | 19.5 | 8 | 8.2 | 135 | 16.0 | 24 | 18.6 | 17 | 13.8 |
| No. of Test Used |  |  |  |  |  |  |  |  |  |  |  |  |  |  |
| *Single* | 1458 | 80.7 | 84 | 94.4 | 432 | 82.8 | 86 | 87.8 | 640 | 75.7 | 118 | 91.5 | 98 | 79.7 |
| *Multiple* | 349 | 19.3 | 5 | 5.6 | 90 | 17.2 | 12 | 12.2 | 206 | 24.3 | 11 | 8.5 | 25 | 20.3 |
| Assay Types |  |  |  |  |  |  |  |  |  |  |  |  |  |  |
| *Commercial ELISA* | 655 | 36.2 | 26 | 29.2 | 174 | 33.3 | 30 | 30.6 | 357 | 42.2 | 39 | 30.2 | 29 | 23.6 |
| *Commercial LFIA* | 308 | 17.0 | 37 | 41.6 | 71 | 13.6 | 13 | 13.3 | 145 | 17.1 | 6 | 4.7 | 36 | 29.3 |
| *Commercial IFA* | 33 | 1.8 | 3 | 3.4 | 11 | 2.1 | 2 | 2.0 | 15 | 1.8 | 0 | 0.0 | 2 | 1.6 |
| *Commercial CLIA* | 879 | 48.6 | 12 | 13.5 | 258 | 49.4 | 47 | 48.0 | 428 | 50.6 | 63 | 48.8 | 71 | 57.7 |
| *Commercial Other^b^* | 14 | 0.8 | 0 | 0.0 | 5 | 1.0 | 0 | 0.0 | 8 | 0.9 | 0 | 0.0 | 1 | 0.8 |
| *Self-developed assay* | 167 | 9.2 | 5 | 5.6 | 31 | 5.9 | 8 | 8.2 | 95 | 11.2 | 2 | 1.6 | 26 | 21.1 |
| *Unknown* | 287 | 15.9 | 3 | 3.4 | 100 | 19.2 | 9 | 9.2 | 128 | 15.1 | 32 | 24.8 | 15 | 12.2 |
| Test-adjusted estimation^c^ |  |  |  |  |  |  |  |  |  |  |  |  |  |  |
| *Not Adjusted* | 1559 | 86.3 | 68 | 76.4 | 451 | 86.4 | 86 | 87.8 | 733 | 86.6 | 108 | 83.7 | 113 | 91.9 |
| *Adjusted* | 248 | 13.7 | 21 | 23.6 | 71 | 13.6 | 12 | 12.2 | 113 | 13.4 | 21 | 16.3 | 10 | 8.1 |

*Abbreviations: ELISA: enzyme-linked immunosorbent assay; LFIA: lateral flow serology assay; IFA: immunofluorescence assays; CLIA: chemiluminescent serology assay; CMIA: chemiluminescent microparticle serology assay; RDT: rapid diagnostic tests.*

*Notes: ^a^ Q1-Q2 was between Jan. 1st and Jun. 30th of the given year; Q3-Q4 was between Jul. 1st and Dec. 31st of the given year.*

*^b^ Other commercial serology assays include Luminex, and other test types were not specified by manufacturers.*

*^c^ Bayesian methods were adopted to adjust for misclassification due to test Sn. / Sp. for more accurate prevalence estimations*

### **Figure S2. Pareto chart of serological assay use.**

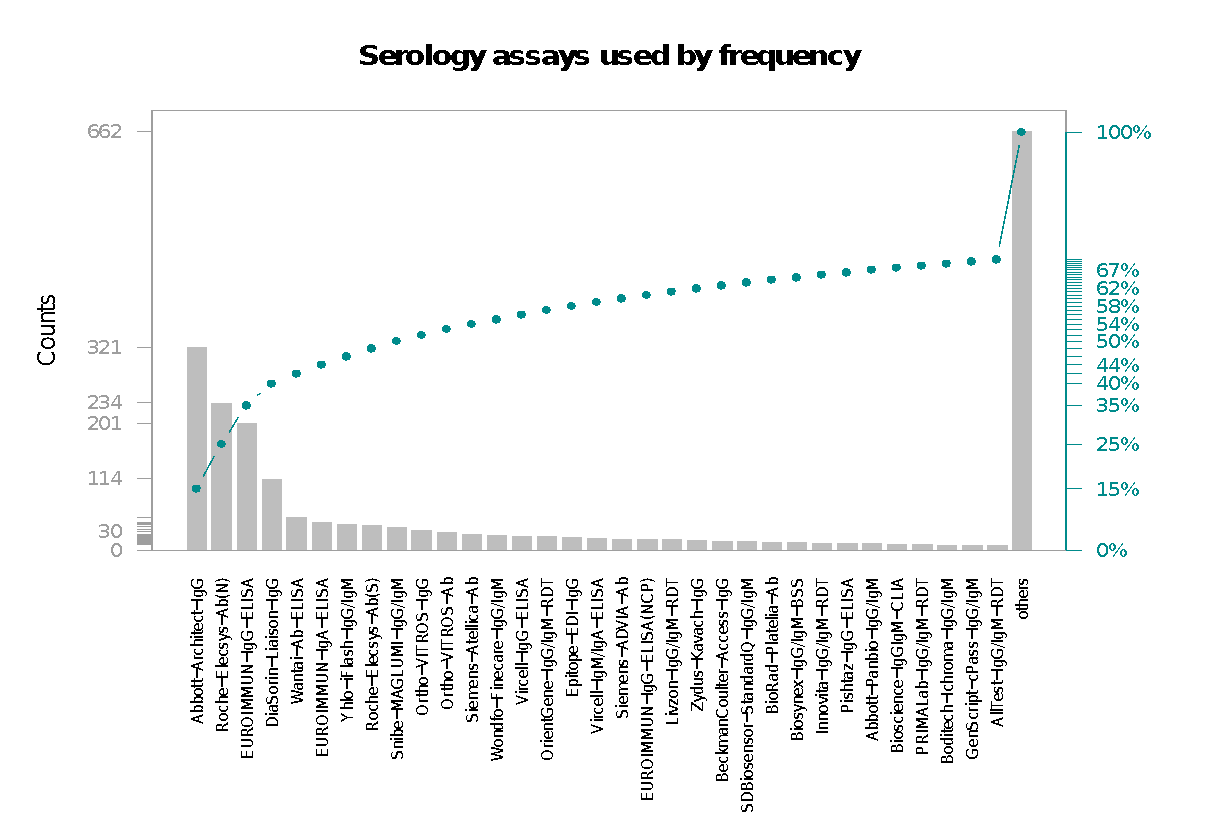

*Note: Plot shows the frequency of serological assay use in 1807 serosurveys. Immunoassays are sorted by frequency of use. The top 25 immunoassays are shown individually with the remainder being grouped as “others”.*

### **Figure S3.1 Difference in reported assay performance among independent evaluation, third party evaluation, and manufacturer evaluation among top 50 assays (ELISA)**

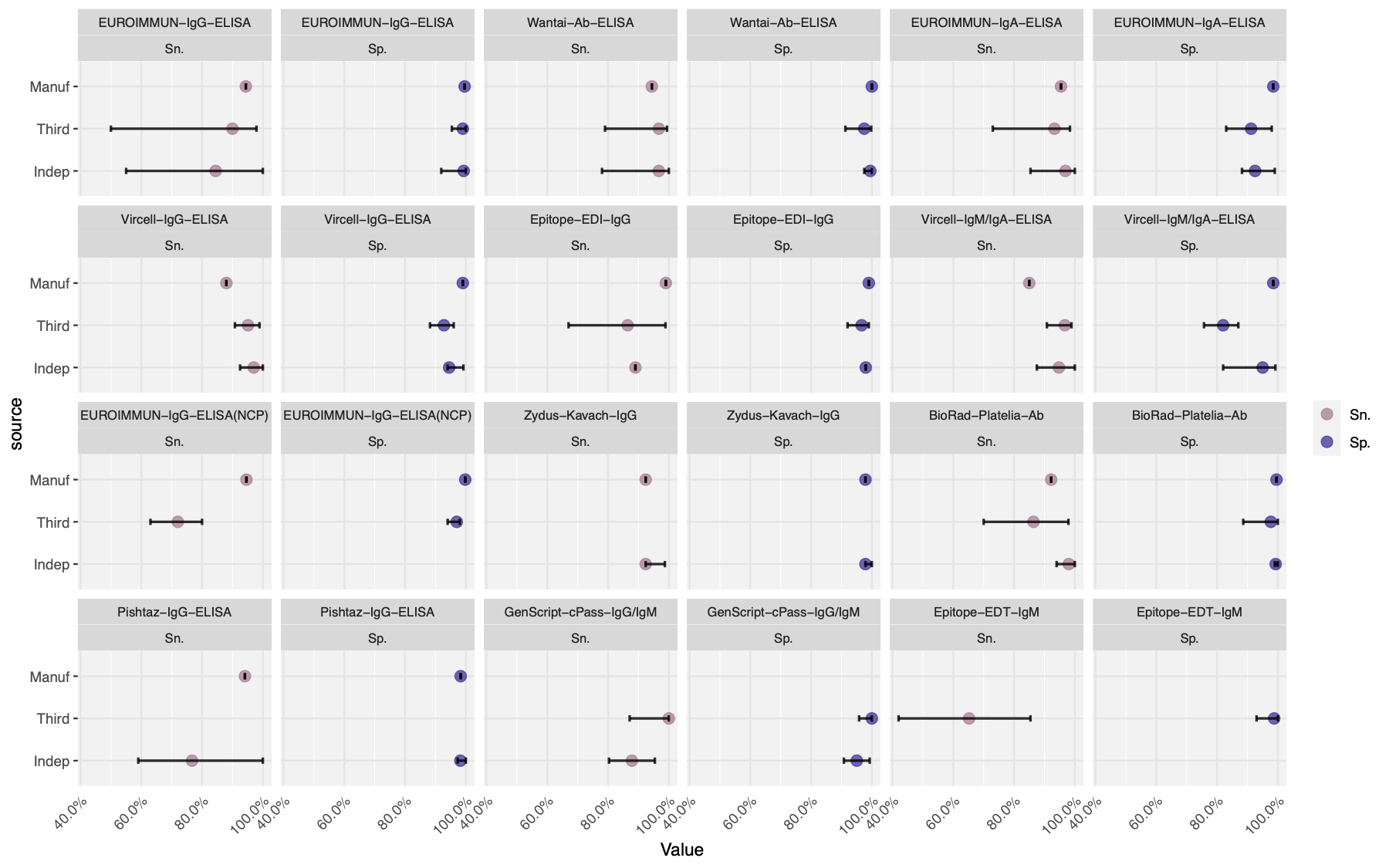

*Note. The figure shows the side-by-side comparison of assay performance for the top 25 assays. Performance evaluations came from three sources: manufacturer reports, third-party labs and independent groups. Intervals were constructed by computing the minimum/maximum performance values from the given source.*

### **Figure S3.2 Difference in reported assay performance among independent evaluation, third party evaluation, and manufacturer evaluation among top 50 assays (CLIA)**

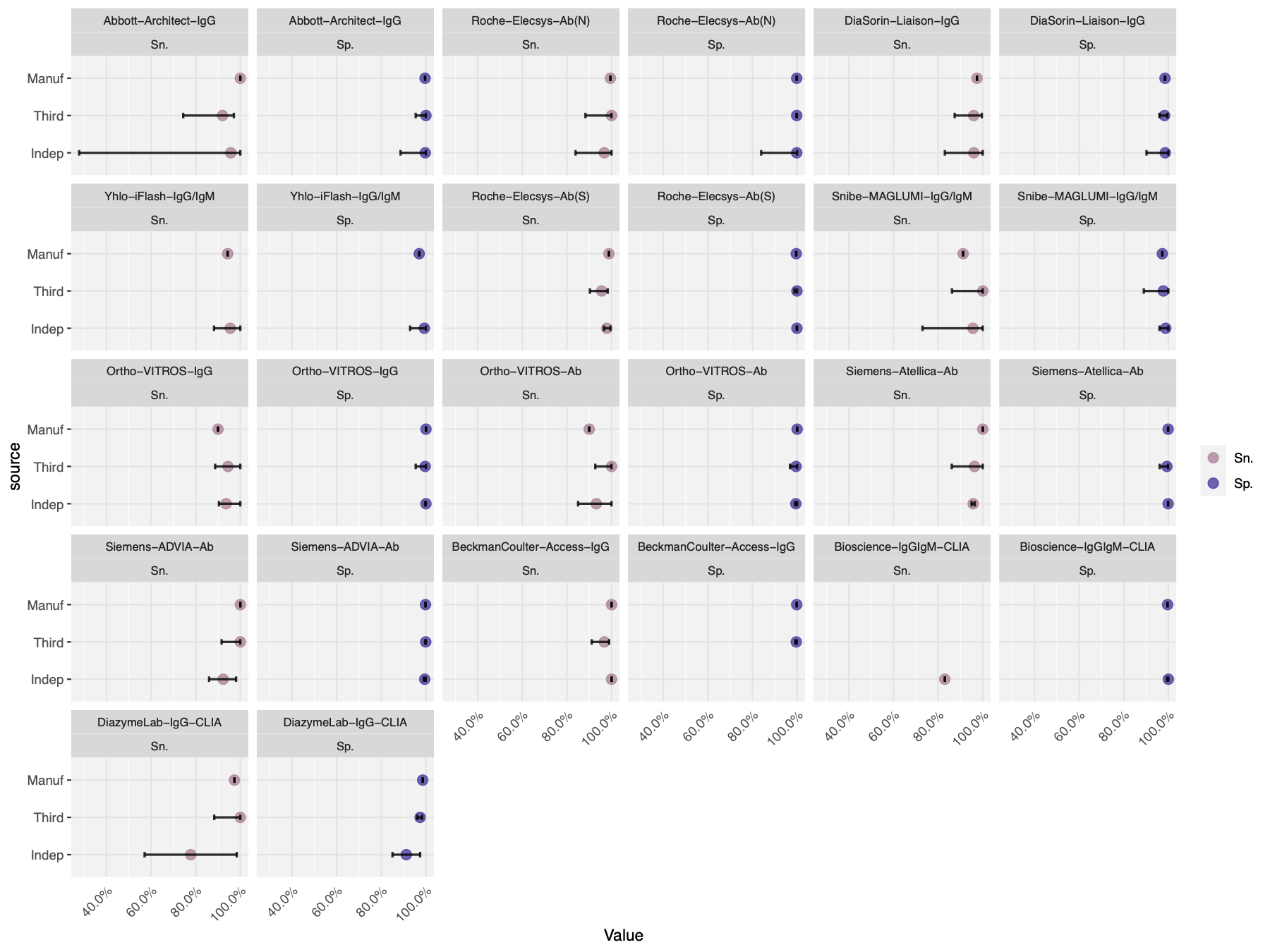

### **Figure S3.3 Difference in reported assay performance among independent evaluation, third party evaluation, and manufacturer evaluation among top 50 assays (LFIA/IFA)**

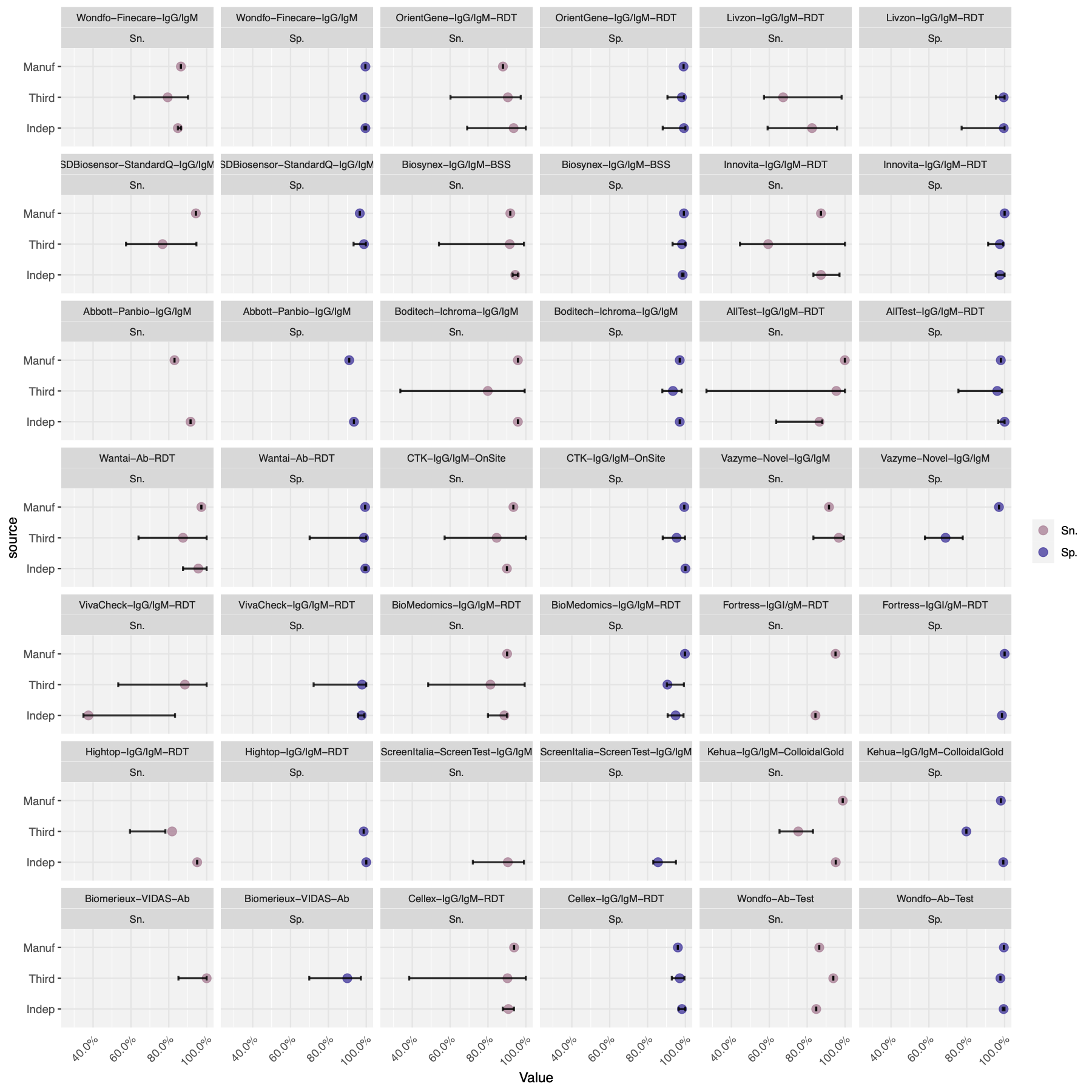

### **Figure S4. Pairwise Bland-Altman plots on the agreement of Sn. and Sp. between a source of assay evaluation against the manufacturer evaluation: Manufacturer vs. (a)-(b) Australia NRL, (c)-(d) US FDA, (e)-(f) Netherland CIDC, (g)-(h) FIND Diagnostics, (i)-(j) Australia Doherty, (k)-(l) independent evaluations**

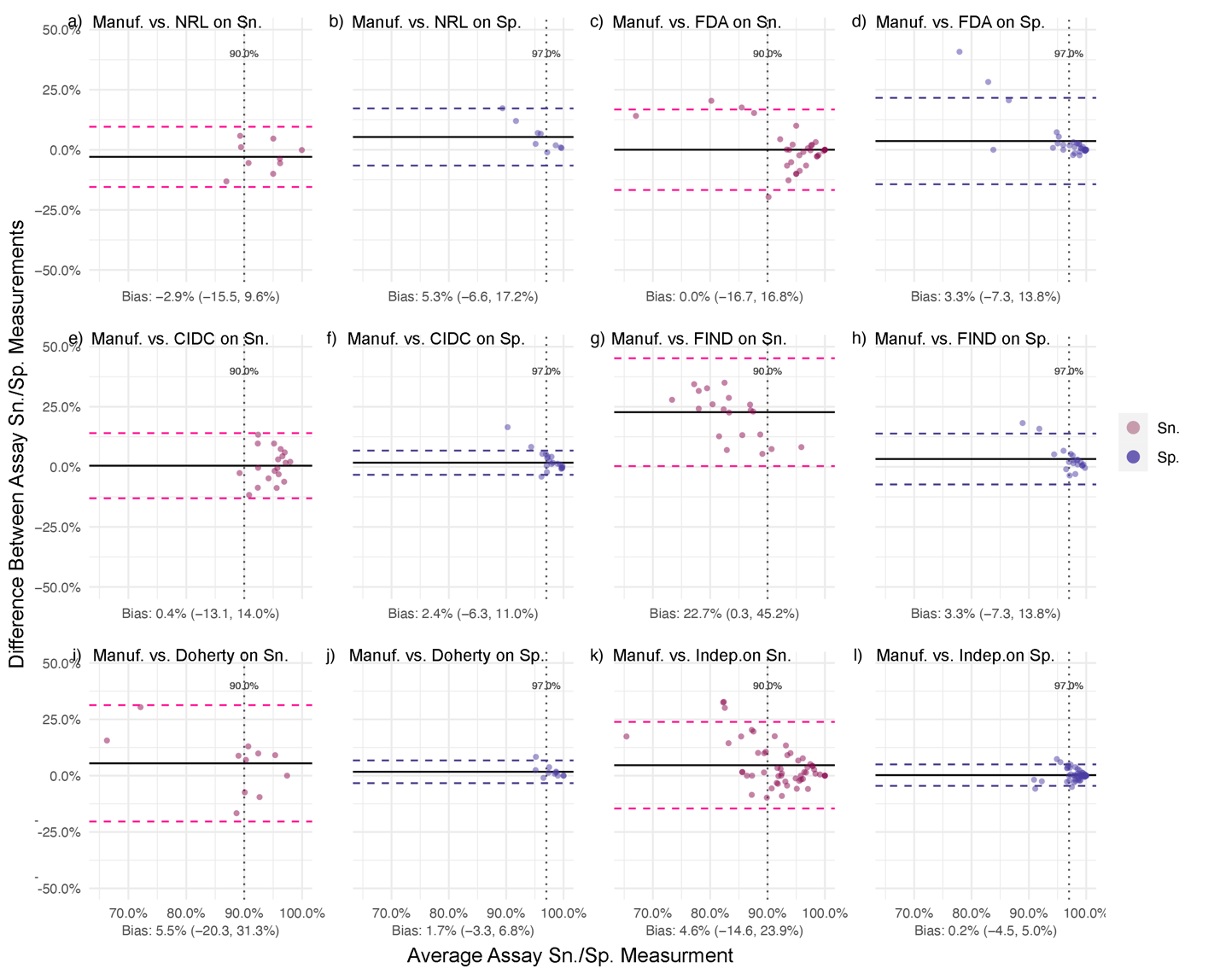

*Note. Assays missing corresponding data were not involved in the analysis. The bias level (with its 95% confidence interval) was the average difference given by performance value from manufacturers - performance value from the other evaluation source.*

### **Figure S5. Pairwise Bland-Altman plots on the agreement of Sn. and Sp. between a source of lab evaluation against independent group evaluations: Independent vs. (a)-(b) Australia NRL, (c)-(d) US FDA, (e)-(f) Netherland CIDC, (g)-(h) FIND Diagnostics, (i)-(j) Australia Doherty.**

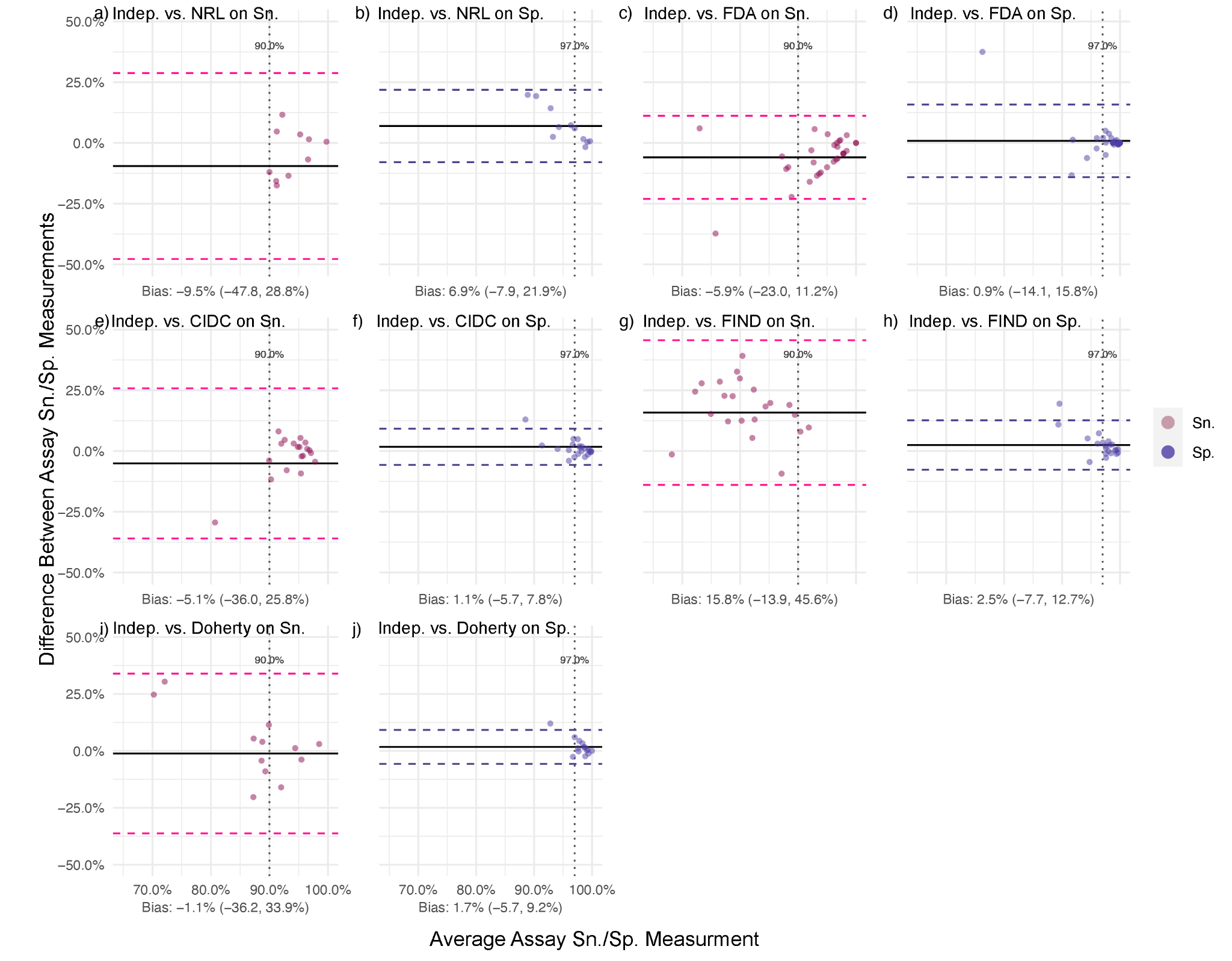

*Note. Assays missing corresponding data were not involved in the analysis. The bias level (with its 95% confidence interval) was the average difference given by performance value from manufacturers - performance value from the other evaluation source*

### **Table S4. Categorizing the top 50 assays based on the median performance value given by** **third-party and independent evaluations**

| **Group** | **A**  **(Sn: 95.0-99.9%,**  **Sp: 97.0-99.9%;**  **n = 10)** | **B**  **(Sn: 90.0-95.0%, Sp: 90.0-97.0%;**  **n = 8)** | **C**  **(Sn: 80.0-90.0%,**  **Sp: 87.0-90.0%;**  **n = 18)** | **D**  **(Sn: <80.0%,**  **Sp: <87.0%;**  **n = 18)** |
| --- | --- | --- | --- | --- |
| **Serological assays (Abbv.)** | Abbott-Architect-IgG, Roche-Elecsys-Ab(N), Yhlo-iFlash-IgG/IgM, GenScript-cPass-IgG/IgM, Roche-Elecsys-Ab(S), Wantai-Ab-ELISA, Siemens-Atellica-Ab, Ortho-VITROS-Ab, Siemens-ADVIA-Ab, BeckmanCoulter-Access-IgG | DiaSorin-Liaison-IgG, BioRad-Platelia-Ab, Snibe-MAGLUMI-IgG/IgM, PRIMALab-IgG/IgM-RDT, Ortho-VITROS-IgG, OrientGene-IgG/IgM-RDT, Biosynex-IgG/IgM-BSS, Wantai-Ab-RDT | EUROIMMUN-IgG-ELISA, Boditech-Ichroma-IgG/IgM, Bioscience-IgGIgM-CLIA, EUROIMMUN-IgA-ELISA, Cellex-IgG/IgM-RDT, Vircell-IgG-ELISA, Vircell-IgM/IgA-ELISA, Abbott-Panbio-IgG/IgM, Epitope-EDI-IgG, BioMedomics-IgG/IgM-RDT, Wondfo-Finecare-IgG/IgM, DiazymeLab-IgG-CLIA, CTK-IgG/IgM-OnSite, Biomerieux-VIDAS-Ab, Wondfo-Ab-Test, Fortress-IgGI/gM-RDT, Kehua-IgG/IgM-ColloidalGold, Hightop-IgG/IgM-RDT, Zydus-Kavach-IgG | AllTest-IgG/IgM-RDT, Innovita-IgG/IgM-RDT, Pishtaz-IgG-ELISA, VivaCheck-IgG/IgM-RDT, Livzon-IgG/IgM-RDT, EUROIMMUN-IgG-ELISA(NCP), Vazyme-Novel-IgG/IgM, ScreenItalia-ScreenTest-IgG/IgM, Epitope-EDT-IgM, SDBiosensor-StandardQ-IgG/IgM |

### **Table S5. Testing algorithms involving multiple serological assays**

| **Multiple Test Combination** | **Studies** | |
| --- | --- | --- |
|  | **(N = 349)** | |
|  | **n** | **%** |
| No. of tests used |  |  |
| *Two tests* | 281 | 80.5 |
| *Three or more* | 68 | 19.5 |
| Test types combined |  |  |
| *Commercial & Commercial* | 28 | 8.0 |
| *Commercial & Self-developed* | 67 | 19.2 |
| *Self-developed & Self-developed* | 254 | 72.8 |
| Assay modalities combined |  |  |
| *RDT with RDT* | 20 | 5.7 |
| *RDT with Non-RDT (*)* | 82 | 23.5 |
| *Non-RDT with Non-RDT* | 267 | 76.5 |
| Antibody targets combined |  |  |
| *S-Test with S-Test* | 121 | 34.7 |
| *S-Test with N-Test* | 152 | 43.6 |
| *Other/ Unknown* | 76 | 21.8 |
| Combined Sn. and Sp. |  |  |
| *Reported* | 33 | 9.5 |
| *Not reported* | 283 | 81.1 |
| Testing Algorithm |  |  |
| *Participants considered seropositive if at least one test is positive (Parallel OR)* | 101 | 28.9 |
| *Participants considered seropositive if all tests are positive (Parallel AND)* | 47 | 13.5 |
| *One test on all participants, with a second test used to confirm positives; participants considered seropositive if both tests were positive (Series AND)* | 111 | 31.8 |
| *Different singular tests (non-overlapping testing) used on different participants* | 30 | 8.6 |
| *Unclear* | 60 | 17.2 |

*Note: * Non-RDT tests included ELISA, CLIA (including CGIA and CGIA), and neutralization assays*

### **Figure S6. Bland-Altman plot on the agreement between serological testing results with multiple assays and with a single assay: 1) any single assay types (N = 167), 2) single assay: S-tests, 3) single assay: N-tests, 4) single assay: multiplexed tests, 5) single assay: RDTs, 6) single assay: ELISAs, 7) single assay: CLIAs, 8) single assay: LFIA/IFAs**

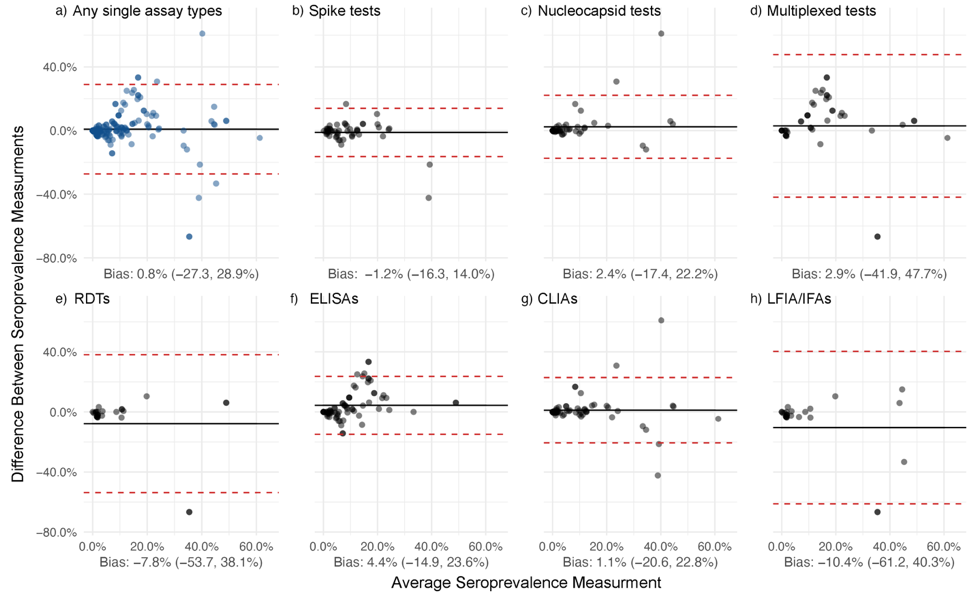

*Note. Assays missing corresponding data were not involved in the analysis. The bias level (with 95% confidence interval) was the average difference given by seroprevalence estimated by the multiple testing - seroprevalence estimated by single testing.*

### **Table S6. List of identified commercial serology assays from systematic review**

| No. | Manufacturer | Test Name | Country | Type | Isotype | Antibody Target | Sn.  (Manuf., %) | Sp.  (Manuf., %) | Sn.^[1]^  (Third., %) | Sp.  (Third., %) | Sn.^[2]^  (Indep., %) | Sp.  (Indep., %) |
| --- | --- | --- | --- | --- | --- | --- | --- | --- | --- | --- | --- | --- |
| 1 | AAZ LMB | COVID-PRESTO | France | LFIA | IgG,IgM | Nucleocapsid | 100.0 | 100.0 |  |  | 100.0  (100.0, 100.0) | 100.0  (100.0, 100.0) |
| 2 | Abbexa | Abbexa COVID-19 IgG/IgM Rapid Test Kit | UK | LFIA | IgG,IgM | Spike | 98.5 | 97.9 | 96.7  (83.3, 99.4) | 96.7  (83.3, 99.4) | - | - |
| 3 | Abbott Laboratories | Abbott Architect SARS-CoV-2 IgG | USA | CLIA | IgG | Nucleocapsid | 100.0 | 99.6 | 92  (74.4, 97.1) | 92  (74.4, 97.1) | 95.7  (27.8, 100.0) | 99.7  (88.6, 100.0) |
| 4 | Abbott Laboratories | AdviseDX SARS-CoV-2 anti-IgG II assay | USA | CLIA | IgG | Spike | - | - | - | - | 98.1  (98.1, 98.1) | 99.6  (99.6, 99.6) |
| 5 | Abbott Laboratories | SARS-CoV-2 IgG II Quant | USA | CLIA | IgG | Spike | - | - | - | - | - | - |
| 6 | Abbott Laboratories | Panbio COVID-19 IgG/IgM rapid test device | USA | LFIA | IgG,IgM | Nucleocapsid | 83.0 | 91.0 | - | - | 91.5  (91.5, 91.5) | 93.5  (93.5, 93.5) |
| 7 | Abbott Laboratories | Architect Anti-SARS-CoV-2 IgM | USA | CLIA | others | Spike | - | - | - | - | - | - |
| 8 | Abnova | COVID-19 Human IgM IgG Assay Kit | China | ELISA | IgG,IgM | NA | 94.4 | 98.5 | - | - | - | - |
| 9 | Academia Sinica | Academia Sinica ELISA assay | China | ELISA | IgG,IgM | Spike | - | - | - | - | - | - |
| 10 | ACRO Biotech Inc | Acro Biotech COVID-19 IgM/IgG Rapid POC test | USA | LFIA | IgG,IgM | NA | - | - | 91.9  (78.7, 97.2) | 91.9  (78.7, 97.2) | 88.0  (37.0, 96.0) | 94.0  (79.2, 99.0) |
| 11 | Alfa Scientific Designs | Instant-view IgG/IgM Antibody COVID- 19 test | USA | LFIA | IgG,IgM | NA | 97.8 | 94.6 | 100.0  (88.7, 100.0) | 100.0  (88.7, 100.0) | - | - |
| 12 | AMEDA Labordiagnostik GmbH | AMP Rapid Test SARS-CoV-2 IgG/IgM | Austria | LFIA | IgG,IgM | NA | 91.8 | 96.4 | - | - | 92.0  (92.0, 92.0) | 99.4  (99.4, 99.4) |
| 13 | AnshLabs | SARS-CoV-2 IgG ELISA | USA | ELISA | IgG | Multiplex targets | - | - | - | - | - | - |
| 14 | Artron Laboratories Inc. | One Step Novel Coronavirus (COVID-19) IgM/IgG Antibody Test | Canada | LFIA | IgG,IgM | NA | 93.4 | 97.7 | 96.2 (87.8, 98.2) | 96.2 (87.8, 98.2) | 83.3  (83.3, 83.3) | 100.0  (100.0, 100.0) |
| 15 | Assure Tech. (Hangzhou) Co., Ltd | COVID-19 IgG/IgM Rapid Test Device | China | LFIA | IgG,IgM | Multiplex targets | - | - | 99.5  (50.9, 100.0) | 99.5  (50.9, 100.0) | 100.0  (100.0, 100.0) | 98.8  (98.8, 98.8) |
| 16 | Aurora Biomed Inc | COVID-19 IgG/IgM Rapid Test | Canada | LFIA | IgG,IgM | NA | - | - | 76.7  (59.1, 88.2) | 76.7  (59.1, 88.2) | - | - |
| 17 | Baiya Phytopharm | Baiya Rapid COVID-19 IgG/IgM Test Kit | Thailand | LFIA | IgG,IgM | Multiplex targets | - | - | - | - | 94.1  (88.7, 94.1) | 98.0  (90.63, 98.0) |
| 18 | Beckman Coulter | Access SARS-CoV-2 IgG Antibody Test | USA | CLIA | IgG | Spike | 100.0 | 99.8 | 96.8  (91.1, 98.9) | 96.8  (91.1, 98.9) | 100.0  (100.0, 100.0) | - |
| 19 | Beijing Hotgen Biotech Co. Ltd, China | COVID-19 IgM/IgG Total Antibody Rapid Test | China | NA | IgG,IgM | NA | - | - | - | - | - | - |
| 20 | Beijing Lepu Medical Technology | Leccurate SARS-CoV-2 Antibody Rapid Test Kit (Colloidal Gold Immunochromatography) | China | LFIA | IgG,IgM | Nucleocapsid | 98.8 | 97.6 | - | - | 66.0  (66.0, 66.0) | 97.0  (97.0, 97.0) |
| 21 | Beijing Wantai Biological | Wantai SARS-CoV-2 IgG ELISA | China | ELISA | IgG | NA | - | - | 100.0  (100.0, 97.64) | 100.0  (100.0, 97.64) | 82.5  (82.5, 82.5) | 98.0  (98.0, 98.0) |
| 22 | Beijing Wantai Biological | Wantai Rapid Test for Total Antibody to SARS-CoV-2 | China | LFIA | IgG,IgM,IgA | Spike | 97.2 | 99.4 |  |  | 95.6  (87.5, 100.0) | 99.5  (99.1, 100.0) |
| 23 | Beijing Wantai Biological | Wantai SARS-CoV-2 Total Ab ELISA | China | ELISA | IgG,IgM,IgA | Spike | 94.4 | 100.0 | 96.7  (79.0, 99.4) | 96.7  (79.0, 99.4) | 96.7  (78.0, 100.0) | 99.5  (97.5, 100.0) |
| 24 | Beijing Wantai Biological | Wantai SARS-CoV-2 IgM ELISA | China | ELISA | others | Spike | - | - | 86.2 (69.0, 96.3) | 86.2 (69.0, 96.3) | - | - |
| 25 | BGI Genomics | SARS-CoV-2 Virus IgG Antibody Detection Kit (ELISA) | China | ELISA | IgG | NA | 98.7 | 98.4 | - | - | - | - |
| 26 | Bio-rad | Platelia SARS-CoV-2 Total Ab assay | USA | ELISA | IgG,IgM,IgA | Nucleocapsid | 92.2 | 99.6 | 86.4  (70.0, 97.9) | 86.4  (70.0, 97.9) | 98.0  (94.0, 100.0) | 99.3  (99.0, 100.0) |
| 27 | Biohit Health Care Ltd | BIOHIT SARS-CoV-2 antibody test | UK | LFIA | IgG,IgM | NA | - | - | 96.4  (83.3, 99.4) | 96.4  (83.3, 99.4) | 97.5  (97.5, 97.5) | 100.0  (100.0, 100.0) |
| 28 | Biolidics Ltd | 2019 nCoV IgM/IgG detection kit | Singapore | CLIA | IgG,IgM | Spike | - | - | 96.7  (83.3, 99.4) | 96.7  (83.3, 99.4) | - | - |
| 29 | BioM√©rieux | VIDAS SARS-COV-2 IgG test | France | other | IgG | Spike | - | - | 100.0  (88.3, 100.0) | 100.0  (88.3, 100.0) | - | - |
| 30 | BioMedomics, Inc. | COVID-19 IgM-IgG Dual Antibody Rapid Test | USA | LFIA | IgG,IgM | Spike | 90.1 | 99.7 | 81.3  (48.4, 99.4) | 81.3  (48.4, 99.4) | 88.7  (80.0, 90.0) | 94.8 (90.6, 99.0) |
| 31 | Biomerieux | VIDAS Multiparametric immunoassay system for medium throughput | France | FIA | IgG,IgM,IgA | Nucleocapsid | - | - | 100.0  (85.1, 100.0) | 100.0  (85.1, 100.0) | - | - |
| 32 | Bioscience Diagnostics | Magnetic Chemiluminescence Enzyme Immunoassay Kit | China | CLIA | IgG,IgM | Multiplex targets | - | 99.7 | - | - | 83.0  (83.0, 83.0) | 100.0  (99.3, 100.0) |
| 33 | Biosense Technologies, India | Coviscreen kittotal antibody (IgM+IgA+IgG) | India | NA | IgG,IgM,IgA | NA | 100.0 | 99.0 | - | - | - | - |
| 34 | Biosynex | Biosynex COVID-19 BSS assay | France | LFIA | IgG,IgM | Spike | 91.8 | 99.2 | 91.5  (54.1, 99.0) | 91.5  (54.1, 99.0) | 94.4  (93.0, 95.8) | 98.5  (98.1, 99.0) |
| 35 | BioSys Laboratories | The BioSys Plus COVID-19 IgM/IgG Rapid Test | Republic of Korea | LFIA | IgG,IgM | NA | 93.5 | 100.0 | - | - | - | - |
| 36 | Biozek | Biozek Medical COVID-19 Rapid Test | Netherlands | LFIA | IgG,IgM | NA | - | - | 95.4  (90.3, 97.9) | 95.4  (90.3, 97.9) | 66.0  (66.0, 66.0) | 97.0  (97.0, 97.0) |
| 37 | Boditech Med Inc. | AFIAS COVID-19 Ab assay | Republic of Korea | FIA | IgG,IgM | Multiplex targets | 99.1 | 94.1 | 85.7  (48.7, 97.2) | 85.7  (48.7, 97.2) | - | - |
| 38 | Boditech Med Inc. | Ichroma COVID-19 Ab Test | Republic of Korea | FIA | IgG,IgM | NA | 95.8 | 97.0 | 79.9  (33.7, 99.4) | 79.9  (33.7, 99.4) | 95.8  (95.8, 95.8) | 97.0  (97.0, 97.0) |
| 39 | Calbiotech Inc. | ErbaLisa COVID-19 IgG ELISA Kit | USA | ELISA | IgG | NA | 98.3 | 99.0 | - | - | - | - |
| 40 | Cambridge Network | COVID-19 IgG and IgM Rapid Test | NA | LFIA | IgG,IgM | NA | - | - | - | - | 100.0  (100.0, 100.0) | 100.0  (100.0, 100.0) |
| 41 | Celer Technologies Inc. | One Step COVID-19 Test | Brazil | LFIA | IgG,IgM | NA | 86.4 | 97.6 | - | - | 86.4  (86.4, 86.4) | 99.6  (99.6, 99.6) |
| 42 | Cellex Inc. | COVID-19 IgG/IgM Rapid Test Cassette | USA | LFIA | IgG,IgM | Nucleocapsid | 93.8 | 96.0 | 90.3  (38.4, 100.0) | 90.3  (38.4, 100.0) | 90.8  (87.8, 93.8) | 98.2  (96.4, 99.9) |
| 43 | ChemBio Diagnostics Inc. | DPP COVID-19 IgM/IgG System | USA | NA | IgG,IgM | Nucleocapsid | - | - | 82.1  (64.4, 92.1) | 82.1  (64.4, 92.1) | - | - |
| 44 | CHiL | COVID-19 IgG/ IgM Rapid Test Cassette | Turkey | LFIA | IgG,IgM | Multiplex targets | - | - | - | - | - | - |
| 45 | CLUNGENE | CLUNGENE SARS-COV-2 VIRUS (COVID-19) IgG/IgM Rapid Test Cassette lateral flow immunoassay | Germany | LFIA | IgG,IgM | NA | - | - | - | - | - | - |
| 46 | CONICET | COVIDAR IgM and IgG ELISA | Argentina | ELISA | IgG,IgM | NA | - | - | - | - | 90.0  (90.0, 90.0) | 100.0  (100.0, 100.0) |
| 47 | Core Technology Co. Ltd | Coretests COVID-19 IgM/IgG Ab Test | China | LFIA | IgG,IgM | Multiplex targets | 97.6 | 100.0 | 68.9  (57, 78.7) | 68.9  (57, 78.7) | 67.5  (66.0, 69.0) | 97.0  (97.0, 97.0) |
| 48 | COVIDAR | SEROKIT-ELISA COVIDAR IgG | Argentina | ELISA | IgG | NA | 95.0 | 100.0 | - | - | 96.7  (96.7, 96.7) | 100.0  (100.0, 100.0) |
| 49 | CTK Biotech, Inc. | OnSite COVID-19 IgG/IgM Rapid Test | USA | LFIA | IgG,IgM | Spike | 93.4 | 99.4 | 84.6  (57.1, 100.0) | 84.6  (57.1, 100.0) | 90.0  (90.0, 90.0) | 100.0  (100.0, 100.0) |
| 50 | Diagnostic Bioprobes Srl | IgG and IgM anti-SARS-CoV-2 ELISA | Italy | ELISA | IgG,IgM | Multiplex targets | 98.0 | 98.0 | - | - | - | - |
| 51 | DiaSorin SpA | Liaison SARS-CoV-2 S1/S2 IgG | Italy | CLIA | IgG | Spike | 97.4 | 98.5 | 95.9  (87.4, 99.6) | 95.9  (87.4, 99.6) | 96.0  (83.0, 100.0) | 98.6  (90.2, 100.0) |
| 52 | DiaSorin SpA | LIAISON SARS-CoV-2 S1/S2 IgM | Italy | CLIA | others | Spike | - | - | - | - | - | - |
| 53 | DiaSorin SpA | Liason SARS-CoV-2 IgM test | Italy | CLIA | others | Spike | - | - | 91.8  (85.6, 95.5) | 91.8  (85.6, 95.5) | - | - |
| 54 | DIAsource | anti-SARS-CoV-2 IgG ELISA | Belgium | ELISA | IgG | NA | - | - | - | - | - | - |
| 55 | Diazyme Laboratories Inc | Diazyme DZ-Lite SARS-CoV-2 IgG CLIA tests | USA | CLIA | IgG | Multiplex targets | 97.4 | 98.6 | 100.0  (88.3, 100.0) | 100.0  (88.3, 100.0) | 77.8  (57.1, 98.4) | 91.2  (85.0, 97.4) |
| 56 | Diazyme Laboratories Inc | Diazyme DZ-Lite SARS-CoV-2 IgM CLIA tests | USA | CLIA | others | Nucleocapsid | - | - | 94.4  (88.4, 97.4) | 94.4  (88.4, 97.4) | 57.1  (57.1, 57.1) | 85.0  (85.0, 85.0) |
| 57 | Durviz S.L. | REAL COVID19 Rapid test cassette | Spain | LFIA | IgG,IgM | NA | 100.0 | 98.0 | - | - | - | - |
| 58 | Dynamiker Biotechnology (Tianjin) Co., Ltd | 2019-nCoV IgG/IgM Rapid Test | China | LFIA | IgG,IgM | Nucleocapsid | - | - | 70.6  (35.3, 91.8) | 70.6  (35.3, 91.8) | - | - |
| 59 | Eagle Bioscience | Coronavirus COVID-19 IgG ELISA Assay | USA | ELISA | IgG | Nucleocapsid | 100.0 | 100.0 | - | - | - | - |
| 60 | ECO Diagnostica LTDA | COVID-19 IgG/IgM ECO Test | Brazil | CLIA | IgG,IgM | Nucleocapsid | 94.5 | 95.7 | - | - | - | - |
| 61 | EKVITESTLAB LLC | EQUI SARS-CoV-2 IgG | Ukraine | ELISA | IgG | NA | - | - | - | - | - | - |
| 62 | EKVITESTLAB LLC | EQUI SARS-CoV-2 IgM | Ukraine | ELISA | others | NA | - | - | - | - | - | - |
| 63 | Enzo Life Sciences | SARS-CoV-2 IgG ELISA | USA | ELISA | IgG | Spike | - | - | - | - | - | - |
| 64 | Epitope Diagnostics, Inc. | EDI‚ Novel Coronavirus COVID-19 IgG ELISA Kit | USA | ELISA | IgG | Nucleocapsid | 99.0 | 99.0 | 86.5  (67, 98.9) | 86.5  (67, 98.9) | 89.0  (89.0, 89.0) | 98.0  (98.0, 98.0) |
| 65 | Epitope Diagnostics, Inc. | EDI‚ Novel Coronavirus COVID-19 IgM ELISA Kit | USA | ELISA | others | NA | - | - | 65.2  (42.0, 85.4) | 65.2  (42.0, 85.4) | - | - |
| 66 | Erba Manneim | ErbaLisa COVID-19 IgG semi quantitative kit | Germany | ELISA | IgG | Multiplex targets | - | - | - | - | - | - |
| 67 | ET Healthcare | Cyclic Enhanced Fluorescence Assay (CEFA) | USA | other | IgG,IgM | NA | - | - | - | - | - | - |
| 68 | ET Healthcare | Pylon COVID-19 IgG/IgM assay | USA | NA | IgG,IgM | Spike | - | - | - | - | 89.0  (89.0, 89.0) | 100.0  (100.0, 100.0) |
| 69 | EUROIMMUN AG | SARS-CoV-2 NCP- IgM | Germany | ELISA | others | Nucleocapsid | - | - | - | - | - | - |
| 70 | EUROIMMUN AG | Anti-SARS-CoV-2 ELISA (IgA) | Germany | ELISA | others | Spike | 95.5 | 98.5 | 93.3  (73, 98.4) | 93.3  (73, 98.4) | 96.9  (85.4, 100.0) | 92.5  (88.2, 99.0) |
| 71 | EUROIMMUN AG | Anti-SARS-CoV-2 ELISA (IgG) | Germany | ELISA | IgG | Spike | 94.4 | 99.6 | 90.0  (50.0, 97.9) | 90.0  (50.0, 97.9) | 84.5  (55.0, 100.0) | 99.3  (91.9, 100.0) |
| 72 | EUROIMMUN AG | SARS-CoV-2-NCP-IgG ELISA | Germany | ELISA | IgG | Nucleocapsid | 94.6 | 99.8 | 72.0  (63.0, 80.0) | 72.0  (63.0, 80.0) | - | - |
| 73 | Exsera BioLabs | Not available | USA | NA | IgG | Nucleocapsid | 85.0 | 98.6 | - | - | - | - |
| 74 | FBSI State scientific center for applied microbiology and biotechnology | ELISA anti-SARS-CoV reagent kit -2 IgG | Russian Federation | ELISA | IgG | Nucleocapsid | - | - | - | - | - | - |
| 75 | Fortress | SARS-CoV-2 ELISA | UK | ELISA | IgG,IgM,IgA | Spike | - | - | - | - | - | - |
| 76 | Fortress Diagnostics | Coronavirus Antibody Rapid Test | UK | LFIA | IgG,IgM | Spike | 95.0 | 100.0 | - | - | 84.4  (84.4, 84.4) | 98.6  (98.6, 98.6) |
| 77 | Genalyte (San Diego, CA) | Maverick Multi-Antigen Serology | USA | other | IgG,IgM | Multiplex targets | - | - | 96.1  (92.2, 98.1) | 96.1  (92.2, 98.1) | - | - |
| 78 | GenBody Inc. | GenBody COVID-19 IgM/IgG | Republic of Korea | LFIA | IgG,IgM | NA | 74.1 | 100.0 | 58.5  (11.0, 75.4) | 58.5  (11.0, 75.4) | 56.7  (40.0, 60.0) | 98.8  (98.8, 100.0) |
| 79 | Genetico | CoronaPass total antibodies test | Russian Federation | ELISA | IgG,IgM,IgA | Spike | 98.7 | 100.0 | - | - | 92.0  (92.0, 92.0) | 100.0  (100.0, 100.0) |
| 80 | Genobio Pharmaceutical (Shanghai China) | Virusee Immunochromatographic Rapid Method | China | LFIA | IgG,IgM | NA | 95.3 | 96.8 | 80.0  (62.7, 90.5) | 80.0  (62.7, 90.5) | - | - |
| 81 | Genrui Biotech | Novel Coronavirus IgG/IgM test kit | China | LFIA | IgG,IgM | NA | 91.0 | 95.4 | - | - | - | - |
| 82 | GenScript | cPass SARS-CoV-2 Surrogate Virus Neutralization Test ELISA Kit | USA | ELISA | IgG,IgM | Spike | - | - | 100.0  (87.1, 100.0) | 100.0  (87.1, 100.0) | 87.9  (80.3, 95.4) | 95.1  (90.8, 99.3) |
| 83 | Getz Pharma | IgG/IgM Test Kit (Colloidal gold) | Pakistan | NA | IgG,IgM | NA | - | - | - | - | - | - |
| 84 | Gold Standard Diagnostic | SARS-CoV-2 IgG ELISA Assay | USA | ELISA | IgG | Nucleocapsid | - | - | - | - | - | - |
| 85 | Guangzhou Wondfo Biotech Co., Ltd | Finecare SARS-CoV-2 Antibody test | China | LFIA | IgG,IgM | Spike | 86.4 | 99.6 | 79.4  (61.8, 90.2) | 79.4  (61.8, 90.2) | 84.8  (84.8, 86.4) | 99.6  (99.0, 99.9) |
| 86 | Guangzhou Wondfo Biotech Co.Ôºå Ltd | Wondfo SARS-CoV-2 Antibody Test | China | LFIA | IgG,IgM,IgA | NA | 86.4 | 99.6 | 93.8  (93.8, 93.8) | 93.8  (93.8, 93.8) | 84.8  (84.8, 84.8) | 99.5  (99.0, 99.9) |
| 87 | Hanghzhou AllTest Biotech Co., Ltd | 2019-nCoV IgG/IgM Rapid Test Cassette | China | LFIA | IgG,IgM | Nucleocapsid | 99.9 | 98.0 | 95.4  (26.8, 100.0) | 95.4  (26.8, 100.0) | 86.5  (63.6, 88.0) | 100.0  (96.6, 100.0) |
| 88 | Hangzhou Biotest Biotech Co., Ltd | RightSign COVID-19 IgG/IgM Rapid Test Cassette | China | LFIA | IgG,IgM | Spike | - | - | 98.0  (58.6, 100.0) | 98.0  (58.6, 100.0) | 84.0  (79.5, 95.0) | 100.0  (100.0, 100.0) |
| 89 | Hangzhou Clongene Biotech Co., Ltd | Lungene 2019-nCoV IgG/IgM Rapid Test | China | LFIA | IgG,IgM | NA | 97.4 | 98.9 | 97.4  (97.4, 97.4) | 97.4  (97.4, 97.4) | 77.1  (77.1, 77.1) | 95.4  (95.4, 95.4) |
| 90 | Hangzhou Laihe Biotech Co., Ltd | LYHER Novel Coronavirus (2019-nCoV) IgM/IgG Antibody Combo Test Kit(Colloidal Gold) | China | LFIA | IgG,IgM | Spike | - | - | 98.5  (88.7, 100.0) | 98.5  (88.7, 100.0) | 100.0  (100.0, 100.0) | 98.8  (98.8, 98.8) |
| 91 | Hangzhou Realy Tech Co., Ltd | Realy-Tech 2019 nCOV/COVID-19 IgG/IgM Rapid Test | China | LFIA | IgG,IgM | Spike | 97.4 | 96.4 | 92.7  (83.3, 99.4) | 92.7  (83.3, 99.4) | - | - |
| 92 | Hangzhou Testsea Biotechnology Co., Ltd | SARS-COV-2-IgG/IgM Test Cassette | China | LFIA | IgG,IgM | Spike | 92.0 | 100.0 | - | - | - | - |
| 93 | Healgen | COVID-19 IgG/IgM Rapid Test Cassette | USA | LFIA | IgG,IgM | Spike | - | - | 100.0  (88.7, 100.0) | 100.0  (88.7, 100.0) | - | - |
| 94 | ID.Vet | ID Screen | France | ELISA | IgG | Nucleocapsid | - | - | - | - | 92.6  (92.6, 92.6) | 99.6  (99.6, 99.6) |
| 95 | IDEAL TASHKHIS Atieh Co | anti-SARS-CoV-2 IgG and IgM ELISA kit | Iran (Islamic Republic of) | ELISA | IgG,IgM | NA | - | - | - | - | 82.0  (82.0, 82.0) | - |
| 96 | In3diagnostic | ERADIKIT‚Ñ¢ COVID19-IgG Indirect ELISA for the detection of anti SARS-CoV2 | Italy | ELISA | IgG | NA | - | - | - | - | - | - |
| 97 | InBios International, Inc. | anti-SARS-CoV-2 IgM and IgG | USA | ELISA | IgG | Spike | - | - | 100.0  (88.7, 100.0) | 100.0  (88.7, 100.0) | - | - |
| 98 | InBios International, Inc. | anti-SARS-CoV-2 IgM | USA | ELISA | others | Spike | - | - | 96.7  (83.3, 99.4) | 96.7  (83.3, 99.4) | - | - |
| 99 | Ingenasa, Eurofins | INgezim COVID 19 DR test | Spain | ELISA | IgG,IgM,IgA | Nucleocapsid | 93.0 | 95.0 | - | - | 92.0  (92.0, 92.0) | 100.0  (100.0, 100.0) |
| 100 | Innovita Biological Technology Co. Ltd | 2019-nCoV IgG Rapid Test | China | LFIA | IgG | NA | 87.3 | 100.0 | - | - | - | - |
| 101 | Innovita Biological Technology Co. Ltd | 2019-nCoV Ab Test (Colloidal Gold) (IgM/IgG Whole Blood/Serum/Plasma Combo) | China | LFIA | IgG,IgM | NA | 87.3 | 100.0 | 59.4  (44.5, 100.0) | 59.4  (44.5, 100.0) | 87.3  (83.3, 97.1) | 97.6  (95.3, 100.0) |
| 102 | Institut Virion/Serion | SERION ELISA agile SARS-CoV-2 | Germany | ELISA | IgG | Spike | - | - | - | - | - | - |
| 103 | J. Mitra & Co. Pvt., Ltd | COVID Kawach IgG Microlisa | India | ELISA | IgG | NA | - | - | - | - | - | - |
| 104 | Kantaro Biosciences, LLC | COVID-SeroKlir, Kantaro Semi-Quantitative SARS-CoV-2 IgG Antibody Kit | USA | ELISA | IgG | Spike | - | - | 99.1  (97.0, 99.8) | 99.1  (97.0, 99.8) | - | - |
| 105 | Karma Azma Andish Co. | COVID-19 IgM/IgG rapid test | Iran (Islamic Republic of) | LFIA | IgG,IgM | NA | - | - | - | - | 71.1  (71.1, 71.1) | 58.7  (58.7, 58.7) |
| 106 | Kovalent | KOVID Ab (COVID-19 IgG / IgM) kit | Sweden | NA | IgG,IgM | NA | 95.8 | 97.0 | - | - | - | - |
| 107 | Kurabo Industries Ltd | SARS-CoV-2 antibody detection kit (IgM/IgG) | Japan | NA | IgG,IgM | NA | 76.4 | 100.0 | - | - | - | - |
| 108 | LakePharma Inc. | SARS-CoV-2 spike protein IgG MMIA assay | USA | other | IgG | Spike | - | - | - | - | 98.0  (98.0, 98.0) | 100.0  (100.0, 100.0) |
| 109 | moLab GmbH | moscreen 2019-NCOV Corona Virus Test | Germany | LFIA | IgG,IgM | NA | - | - | - | - | 98.8  (80.0, 100.0) | 98.8  (97.0, 100.0) |
| 110 | Maccura Biotechnology Ltd | SARS CoV-2 IgM/IgG Antibody Assay Colloidal Gold Complex test | China | LFIA | IgG,IgM | Multiplex targets | - | - | - | - | - | - |
| 111 | Medical Systems | 2019-nCoV IgM/IgG CLIA | China | CLIA | IgG,IgM | NA | - | - | - | - | 25.0  (0.0, 50.0) | 99.0  (98.9, 99.1) |
| 112 | MedLevensohn | MedTest Coronavirus 2019-nCoV IgG/IgM | Spain | LFIA | IgG,IgM | NA | 85.0 | 99.0 | - | - | - | - |
| 113 | MEDsan GmbH | COVID-19 IgG RDT | Germany | LFIA | IgG,IgM | Spike | - | - | 90.0  (74.4, 96.5) | 90.0  (74.4, 96.5) | - | - |
| 114 | Menarini Diagnostics | COVID-19 IgG/IgM Rapid Test Cassette | Italy | LFIA | IgG,IgM | NA | - | 100.0 | - | - | - | - |
| 115 | Meso Scale Discovery | Meso Scale Discovery multiplex assay | USA | NA | IgG | Spike | - | - | - | - | - | - |
| 116 | Mikrogen GmbH | recomLine SARS-CoV-2 IgG | Germany | other | IgG | Multiplex targets | - | - | - | - | - | - |
| 117 | Mikrogen GmbH | recomWell SARS-CoV-2 IgG ELISA | Germany | ELISA | IgG | Nucleocapsid | 98.0 | 98.7 | 96.3  (90.9, 98.6) | 96.3  (90.9, 98.6) | - | - |
| 118 | Mologic | SARS-CoV-2 ELISA Serology | UK | ELISA | IgG | NA | - | - | - | - | - | - |
| 119 | MP Biomedical | Rapid 2019-nCoV IgG/IgM Combo Diagnostic Test Cards | USA | LFIA | IgG,IgM | Multiplex targets | - | - | 96.7  (83.3, 99.4) | 96.7  (83.3, 99.4) | - | - |
| 120 | Multi-G B.V. | COVID-19 IgG/IgM Ab Test Cassette (Whole Blood/Serum/Plasma) | Belgium | LFIA | IgG,IgM | Nucleocapsid | - | - | - | - | 92.2  (92.2, 92.2) | 97.0  (97.0, 97.0) |
| 121 | nal von minden GmbH | NADAL COVID-19 IgG/IgM Test | Germany | LFIA | IgG,IgM | NA | 94.1 | 99.2 | - | - | 100.0  (92.1, 100.0) | 99.0  (97.0, 100.0) |
| 122 | Nanjing Vazyme Medical Technology Co., Ltd | 2019-Novel Coronavirus (2019-nCoV) IgG/IgM Detection Kit | China | LFIA | IgG,IgM | Multiplex targets | 91.5 | 97.0 | 96.7  (83.3, 99.4) | 96.7  (83.3, 99.4) | - | - |
| 123 | NG Biotech Laboratoires | NG-Test finger-prick test | France | LFIA | IgG,IgM | NA | 100.0 | 95.3 | 91.8  (84.5, 95.9) | 91.8  (84.5, 95.9) | 82.5  (82.5, 82.5) | 98.0  (98.0, 98.0) |
| 124 | NovaTec Immundiagnostics GmbH | NovaLisa SARS-CoV-2 IgG | Germany | ELISA | IgG | NA | 100.0 | 100.0 | 77.6  (55.0, 96.7) | 77.6  (55.0, 96.7) | 94.9  (94.9, 94.9) | 96.2  (96.2, 96.2) |
| 125 | NZK Biotech Co., Wuhan, China | dual-target immuno-fluorescence assay | China | LFIA | IgG,IgM | Multiplex targets | 0.0 | 0.0 | - | - | - | - |
| 126 | Omega diagnostics | COVID-19 IgG ELISA | UK | ELISA | IgG | Multiplex targets | 0.0 | 0.0 | - | - | 88.1  (88.1, 88.1) | 93.2  (93.2, 93.2) |
| 127 | Ortho Clinical Diagnostics Inc. | VITROS Immunodiagnostic Products Anti-SARS-CoV-2 IgG | USA | CLIA | IgG | Spike | 90.0 | 100.0 | 94.5  (88.7, 100.0) | 94.5  (88.7, 100.0) | 93.6  (90.4, 100.0) | 100.0  (99.6, 100.0) |
| 128 | Ortho Clinical Diagnostics Inc. | VITROS Immunodiagnostic Products Anti-SARS-CoV-2 Total | USA | CLIA | IgG,IgM,IgA | Spike | 90.0 | 100.0 | 100.0  (92.7, 100.0) | 100.0  (92.7, 100.0) | 93.2  (85, 100.0) | 99.4  (99.0, 100.0) |
| 129 | Palex | Diapro | Spain | ELISA | IgG | Multiplex targets | 98.0 | 90.0 | - | - | 88.9  (88.9, 88.9) | 91.8  (91.8, 91.8) |
| 130 | PCL Inc | PCL COVID19 IgG/IgM Rapid Gold | Republic of Korea | NA | IgG,IgM | NA | - | - | 83.0  (83.3, 99.4) | 83.0  (83.3, 99.4) | - | - |
| 131 | PCL Inc | PCL COVID-19 Total Ab EIA test | Republic of Korea | ELISA | IgG,IgM,IgA | Multiplex targets | 98.2 | 100.0 | - | - | - | - |
| 132 | PerkinElmer | GSP1/DELFIA1 anti SARS-CoV2 kit | Italy | LFIA | IgG | Spike | 96.0 | 100.0 | - | - | - | - |
| 133 | Phamatech | COVID19 RAPID TEST | USA | NA | IgG,IgM | NA | - | - | 86.7  (70.3, 94.7) | 86.7  (70.3, 94.7) | - | - |
| 134 | PharmACT | BELTEST-IT COV-2 Rapid Test | Germany | FIA | IgG,IgM | NA | - | - | - | - | - | - |
| 135 | Pishtaz Diagnostics Iran | ELISA kit Pishtaz Teb Diagnostics | Iran (Islamic Republic of) | ELISA | IgG | Nucleocapsid | 94.1 | 98.3 | - | - | 76.7  (59.0, 100.0) | 98.2  (97.3, 100.0) |
| 136 | Pishtaz Diagnostics Iran | ELISA kit Pishtaz Teb Diagnostics | Iran (Islamic Republic of) | ELISA | others | Nucleocapsid | - | - | - | - | - | - |
| 137 | Polycheck | anti-SARS-CoV-2 IgG immunoblot | Germany | other | IgG | Multiplex targets | - | - | - | - | - | - |
| 138 | Premier Biotech | COVID-19 IgG/IgM Rapid Test Cassette | USA | LFIA | IgG,IgM | NA | - | - | - | - | 84.1  (82.7, 85.6) | 99.5  (99.5, 99.5) |
| 139 | PRIMA Lab S.A. | PRIMA COVID-19 IgG/IgM Rapid Test | Switzerland | NA | IgG,IgM | Multiplex targets | 100.0 | 98.0 | - | - | 100.0  (100.0, 100.0) | 97.8  (96.0, 99.6) |
| 140 | Prognosis Biotech | Rapid Test 2019-nCoV Total Ig | Greece | LFIA | IgG,IgM,IgA | Spike | 98.8 | 100.0 | - | - | - | - |
| 141 | Qingdao Hightop Biotech Co., Ltd | Hightop COVID-19 IgM/IgG Ab Rapid Test Kit | China | LFIA | IgG,IgM | NA | - | - | 81.8  (59.5, 78.2) | 81.8  (59.5, 78.2) | 95.0 (95.0, 95.0) | 100.0  (100.0, 100.0) |
| 142 | Quotient | MosaiQ COVID-19 Antibody Microarray | Switzerland | other | IgG,IgM | Spike | - | - | 93.0  (85.6, 96.8) | 93.0  (85.6, 96.8) | - | - |
| 143 | RayBiotech | Novel Coronavirus IgG antibody test kit (colloidal gold method) | USA | ELISA | IgG | NA | 90.4 | 98.3 | 70.0  (52.1, 83.3) | 70.0  (52.1, 83.3) | 76.0  (76.0, 76.0) | 95.0  (95.0, 95.0) |
| 144 | Roche Diagnostics | Elecsys®Anti‐SARS‐CoV‐2 (N) | Switzerland | CLIA | IgG,IgM,IgA | Nucleocapsid | 99.5 | 99.8 | 100.0  (88.3, 100.0) | 100.0  (88.3, 100.0) | 96.8  (83.9, 100.0) | 99.8  (83.9, 100.0) |
| 145 | Roche Diagnostics | Elecsys®Anti‐SARS‐CoV‐2 (S) | Switzerland | CLIA | IgG,IgM,IgA | Spike | 98.8 | 99.6 | 95.6  (90.3, 98.3) | 95.6  (90.3, 98.3) | 97.9  (96.6, 99.5) | 99.9  (99.8, 99.9) |
| 146 | Salofa Oy manufacturer, Finland | Sienna-Clarity COVIBLOCK COVID-19 IgG/IgM Rapid Test Cassette | Finland | LFIA | IgG,IgM | NA | - | - | 93.3  (78.7, 98.2) | 93.3  (78.7, 98.2) | - | - |
| 147 | Sci LifeLab / KTH | Not available | Sweden | ELISA | NA | Multiplex targets | 99.4 | 98.9 | - | - | 98.9  (98.9, 98.9) | 99.4  (99.4, 99.4) |
| 148 | Screen Italia S.R.L | Screen Test COVID-19 2019-nCOV IgG/IgM | Italy | LFIA | IgG,IgM | NA | - | - | - | - | 90.5  (72.0, 99.0) | 85.5  (83.0, 95.0) |
| 149 | SD Biosensor | Standard Q COVID-19 IgM/IgG Combo | Republic of Korea | FIA | IgG,IgM | NA | 94.5 | 95.7 | - | - | - | - |
| 150 | SD Biosensor | Standard Q COVID-19 IgM/IgG Duo rapid immunochromatography test kit | Republic of Korea | LFIA | IgG,IgM | Nucleocapsid | 94.3 | 96.6 | 76.7  (57.4, 94.6) | 76.7  (57.4, 94.6) | - | - |
| 151 | SeraCare Life Sciences | HRP-conjugated anti human IgM/IgA/IgG | USA | ELISA | IgG,IgM,IgA | Spike | - | - | - | - | - | - |
| 152 | Serimmune | Serum Epitope Repertoire Analysis (SERA) | USA | other | IgG,IgM,IgA | Multiplex targets | 91.0 | 99.3 | - | - | - | - |
| 153 | Shanghai Kehua Bio-engineering Co., Ltd | Diagnostic kit for SARS-CoV-2 IgM/IgG Antibody (Colloidal Gold) | China | LFIA | IgG,IgM | NA | 98.8 | 98.0 | 75.3  (65.4, 83.1) | 75.3  (65.4, 83.1) | 95.1  (95.1, 95.1) | 99.3  (99.3, 99.3) |
| 154 | Shenzhen Mindray Bio-Medical Electronics Co. | Mindray CL-900i anti-SARS-CoV-2 IgG | China | CLIA | IgG | Multiplex targets | - | - | - | - | - | - |
| 155 | Shenzhen Sciarray Biotech Ltd | ELISA test | China | ELISA | IgG,IgM,IgA | Nucleocapsid | - | - | - | - | - | - |
| 156 | Shenzhen Yhlo Biotech Co. Ltd | iFlash-SARS-CoV-2 IgM/IgG | China | CLIA | IgG,IgM | Multiplex targets | 94.3 | 97.0 | - | - | 95.5  (88.2, 100.0) | 99.3  (92.9, 100.0) |
| 157 | Shin Jin Medics Inc | DIAKEY COVID-19 IgM/IgG Rapid Test Kit | Republic of Korea | LFIA | IgG,IgM | Multiplex targets | - | - | - | - | 100.0  (100.0, 100.0) | 92.0  (92.0, 92.0) |
| 158 | Siemens | ADVIA Centaur Immunoassay System | Germany | CLIA | IgG,IgM,IgA | Spike | 100.0 | 99.8 | 100.0  (91.6, 100.0) | 100.0  (91.6, 100.0) | 92.3  (86.0, 98.1) | 99.5  (99.0, 99.9) |
| 159 | Siemens | Atellica IM SARS-CoV-2 Total (COV2T) | Germany | CLIA | IgG,IgM,IgA | Spike | 100.0 | 99.9 | 96.3  (86.1, 100.0) | 96.3  (86.1, 100.0) | 95.7  (95.0, 96.4) | 99.9  (99.9, 99.9) |
| 160 | Snibe Co., Ltd (Shenzhen New Industries Biomedical Engineering Co., Ltd) | MAGLUMI 2019-nCoV IgM/IgG | China | CLIA | IgG,IgM | Multiplex targets | 91.2 | 97.3 | 100.0  (86.2, 100.0) | 100.0  (86.2, 100.0) | 95.6  (73.0, 100.0) | 98.8  (96, 100.0) |
| 161 | Sugentech, Inc. | SGTi-flex COVID-19 IgM/IgG (manual) | Republic of Korea | LFIA | IgG,IgM | NA | - | - | 96.7  (83.3, 99.4) | 96.7  (83.3, 99.4) | - | - |
| 162 | Superbio Biomedical Company | COVID-19 Rapid Test Kit IgG + IgM | China | CLIA | IgG,IgM | NA | 100.0 | 83.8 | 100.0  (88.7, 100.0) | 100.0  (88.7, 100.0) | - | - |
| 163 | SureScreen Diagnostics | COVID-19 antibody rapid test | UK | LFIA | IgG,IgM | Spike | - | - | - | - | 84.4  (74.7, 94.0) | 95.7  (95.1, 96.3) |
| 164 | Synlab | Quantitative IgG antibody test for coronavirus | Germany | CLIA | IgG | Spike | - | - | - | - | - | - |
| 165 | Syntron Bioresearch Inc. | Rapid lgM-lgG Combined Antibody Test for Coronavirus | USA | LFIA | IgG,IgM | Multiplex targets | - | - | - | - | - | - |
| 166 | Sysmex | Nucleocapsid HISCL‚5000/HISCL-800 Automated Immunoassay Systems | Japan | CLIA | IgG | Nucleocapsid | - | - | - | - | - | - |
| 167 | Sysmex | Spike HISCL‚5000/HISCL-800 Automated Immunoassay Systems | Japan | CLIA | IgG | Spike | - | - | - | - | - | - |
| 168 | Tecan | SARS-CoV-2 ELISA IgM and IgG | Switzerland | ELISA | IgG,IgM | NA | - | - | - | - | - | - |
| 169 | Technoclone | 5600100 TECHNOZYM anti SARS-CoV-2 RBD IgG ELISA 96T | Austria | ELISA | IgG | Multiplex targets | - | - | - | - | - | - |
| 170 | Technogenetics srl | TechnoGenetics COVID-19 IgG | Italy | CLIA | IgG | Multiplex targets | 100.0 | 98.6 | - | - | - | - |
| 171 | Technogenetics srl | TechnoGenetics COVID-19 IgM | Italy | CLIA | others | Multiplex targets | 100.0 | 98.6 | - | - | - | - |
| 172 | Tecno Bios srl | SARS-CoV-2 IgG/IgM ELISA Kit IMMUNO-COVID19 | Italy | ELISA | IgG,IgM | Nucleocapsid | - | - | - | - | - | - |
| 173 | TestLine Clinical Diagnostics | Microblot-Array COVID-19 IgA | Czechia | other | others | Multiplex targets | - | - | - | - | - | - |
| 174 | TestLine Clinical Diagnostics | Microblot-Array COVID-19 IgG | Czechia | other | IgG | Multiplex targets | - | - | - | - | - | - |
| 175 | The Binding Site | Human Anti-IgG/A/M SARS-CoV-2 ELISA | UK | ELISA | IgG,IgM,IgA | Spike | - | - | - | - | 98.6  (98.6, 98.6) | 98.3  (98.3, 98.3) |
| 176 | TODA Pharma | TODA Coronadiag + | Japan | LFIA | IgG,IgM | NA | - | - | - | - | - | - |
| 177 | TONYAR Biotech Inc., Taiwan | ASK COVID-19 IgG/IgM Rapid Test | China | LFIA | IgG,IgM | NA | - | - | - | - | - | - |
| 178 | Trinity Biotech | Captia Anti-SARS-CoV-2 (IgG) ELISA | Ireland | ELISA | IgG | Spike | 95.9 | 100.0 | - | - | - | - |
| 179 | Vector BEST | SARS- CoV-2-IgG-EIA-BEST and SARS-CoV-2- IgM-EIA-BEST | Russian Federation | ELISA | IgG,IgM | NA | - | - | - | - | - | - |
| 180 | Vircell S.L. | COVID-19 ELISA IgG | Spain | ELISA | IgG | Multiplex targets | 88.0 | 99.0 | 95.1  (90.8, 98.9) | 95.1  (90.8, 98.9) | 97.0  (92.5, 100.0) | 94.5  (94.0, 99.2) |
| 181 | Vircell S.L. | COVID-19 VIRCLIA IgG MONOTEST | Spain | CLIA | IgG,IgM,IgA | Multiplex targets | - | - | - | - | - | - |
| 182 | Vircell S.L. | COVID-19 ELISA IgM/IgA | Spain | ELISA | others | Multiplex targets | 85.0 | 98.5 | 96.7  (90.8, 98.9) | 96.7  (90.8, 98.9) | 94.8  (87.5, 100.0) | 95.0  (82.0, 99.2) |
| 183 | VivaChek Biotech (Hangzhou) Co., Ltd | VivaDiag COVID-19 IgM/IgG Rapid Test | China | LFIA | IgG,IgM | NA | - | - | 88.5  (53.3, 100.0) | 88.5  (53.3, 100.0) | 37.6  (34.9, 83.3) | 97.6  (95.7, 99.0) |
| 184 | Wuhan UNscience Biotechnology Co., Ltd | UNSCIENCE COVID-19 IgG/IgM antibody Rapid Test Kit | China | LFIA | IgG,IgM | NA | 98.5 | 88.2 | - | - | 66.0  (66.0, 66.0) | 94.0  (94.0, 94.0) |
| 185 | Xiamen Biotime Biotechnology Co., Ltd | SARS-CoV-2 IgG/IgM Rapid Qualitative Test Kit | China | LFIA | IgG,IgM | NA | 80.4 | 99.4 | 97.0  (88.7, 100.0) | 97.0  (88.7, 100.0) | - | - |
| 186 | ZenTech, Angleur, Belgium | QuickZen COVID-19 IgM/IgG kit | Belgium | LFIA | IgG,IgM | Spike | - | - | 55.1  (44.0, 65.1) | 55.1  (44.0, 65.1) | 59.0  (49.2, 68.8) | 100.0  (100.0, 100.0) |
| 187 | ZetaGene Ltd. | ZetaGene COVID-19 Antibody Test IgM/IgG | Sweden | LFIA | IgG,IgM | NA | - | - | - | - | 95.0  (95.0, 95.0) | 97.0  (97.0, 97.0) |
| 188 | Zeus | ELISA SARS-CoV-2 IgG Test System | USA | ELISA | IgG | Nucleocapsid | - | - | 93.3  (78.7, 98.2) | 93.3  (78.7, 98.2) | - | - |
| 189 | Zhejiang Orient Gene Biotech | COVID-19 IgG/IgM Rapid Test | China | LFIA | IgG,IgM | Spike | 87.9 | 99.0 | 90.5  (60.2, 97.3) | 90.5  (60.2, 97.3) | 93.6  (69.0, 100.0) | 99.2  (88.0, 100.0) |
| 190 | Zhuhai Livzon Diagnostics Inc | 2019-nCoV IgG/IgM Antibody Detection Kit | China | LFIA | IgG,IgM | Nucleocapsid | - | - | 67.3  (57.2, 98.2) | 67.3  (57.2, 98.2) | 82.6  (59.1, 95.8) | 99.5  (77.3, 100.0) |
| 191 | Zybio Inc. | SARS-CoV-2 IgM/IgG Antibody Assay Kit | China | LFIA | IgG,IgM | NA | - | - | - | - | - | - |

1. *^,2^ The intervals represent the full ranges of Sn. and Sp. obtained by pooling all performance data from the given evaluation source (i.e., third-party labs, and independent groups). The point estimate is median of the performance value.*
